# The Genomic Landscape of Pediatric Rheumatology Disorders in the Middle East

**DOI:** 10.1101/2020.09.30.20204016

**Authors:** Basil M Fathalla, Ali Alsarhan, Samina Afzal, Maha EL Naofal, Ahmad Abou Tayoun

**Author notes:** Funding Statement: No specific funding or grants supported this work. Conflicts of interest: Authors have no conflicts of interest to disclose. Data Availability: All de-identified data related to this work are presented in the tables and figures. Additional data can be provided upon request. Code Availability: Not applicable. Ethics Statement and consent: Please see Methods.

## Abstract

Genetic investigations for patients with pediatric rheumatological disorders have been limited to classic genotyping testing, mainly *MEFV* hotspot mutation analysis, for periodic fever. Therefore, the landscape and clinical utility of comprehensive genomic investigations for a wider range of pediatric rheumatological disorders have not been fully characterized in the Middle East. Here seventy-one pediatric patients, of diverse Arab origins, were clinically and genetically assessed for a spectrum of rheumatology-related disease at the only dedicated tertiary children’s hospital in the United Arab Emirates. Clinical genomic investigations included mainly (76%) next generation sequencing-based gene panels and whole exome sequencing, along with rapid sequencing in the intensive care unit (ICU) and urgent setting. The overall positive yield was 46.5% (16.7%-66.7% for specific indications), while dual diagnoses were made in 2 cases (3%). Although the majority (21/33, 64%) of positive findings involved the *MEFV* gene, the remaining (12/33, 36%) alterations were attributed to eleven other genes/loci. Copy number variants contributed substantially (5/33, 15.2%) to the overall diagnostic yield. Sequencing-based testing, specifically rapid sequencing, had high positive rate and delivered timely results. Genetic findings guided clinical management plans and interventions in most cases (27/33, 81.8%). We highlight unique findings and provide additional evidence that heterozygous loss of function of the *IFIH1* gene increases susceptibility to recurrent fevers. Our study highlights the importance of comprehensive genomic investigations in patients with pediatric rheumatological disorders, and provides new insights into the pathogenic variation landscape in this group of disorders.

## Introduction

Childhood rheumatic diseases are a heterogeneous group of diseases characterized by chronic inflammation affecting structures of one or more organ systems. Most childhood rheumatic diseases are of unknown cause, however, recent advancements in our knowledge of the functions of the immune system, disease pathology, and the availability of advanced genetic testing had revolutionized our understanding and management of many rheumatic diseases^1,2^. A notable example is the evolution of the concept of autoinflammatory disorders in recent years leading to unveiling many disease mechanisms and pro-inflammatory pathways in ancient diseases such as Familial Mediterranean Fever (FMF) as well as the discovery of an expanding list of newly described monogenic diseases with prominent rheumatological clinical manifestations such as Blau syndrome (NF-kB related disorder) and atypical, severe, early onset rheumatic diseases such as monogenic lupus (Type I interferonopathies)^2,3^. These discoveries also enhanced our ability to manage many of these diseases with targeted therapy such as the use of IL-1 blockers in the cryopyrinopathies^4^.

Another example is the advances in knowledge of the genetic basis of primary immunodeficiencies in which autoimmune disorders may occur. In such cases, patients may present with severe clinical phenotype of known rheumatic diseases, such as sever cutaneous lupus manifestations in complement (C1q) deficiency^5^, and difficult to treat systemic lupus erythematosus (SLE) in purine nucleoside phosphorylase deficiency^6^.

The importance of genetic testing in the rheumatology practice had also increased due to the discovery of the genetic basis of many disorders presenting with non-inflammatory clinical manifestations which mimic rheumatic diseases such as the musculoskeletal manifestations of primary connective tissue diseases and metabolic disorders^7^. For example, the early phase of progressive psuedorheumatoid dysplasia (PPRD) can mimic polyarticular juvenile idiopathic arthritis (polyarticular-JIA) given the polyarticular nature of the disease, prolonged morning stiffness in some patients, and the presence of swollen joints with effusions^7^.

In this study, we report the role of genetic testing in clinical practice in a pediatric rheumatology center in one of the very few tertiary children’s hospitals in the Middle East. Various genetic, primarily sequencing-based, tests were done either to help establish diagnosis, explain unusual contradictory clinical and laboratory manifestations that may result from overlap of more than one disease, or elucidate causes of limited response to medical management. Accordingly, genetic testing was done in the context of 1) work up of suspected autoinflammatory disorders and periodic fever syndromes, 2) patients with difficult to treat inflammatory eye disease, 3) patients with suspected primary noninflammatory connective tissue diseases presenting with musculoskeletal manifestations, and 4) patients with difficult to treat autoimmune connective tissue diseases especially in presence of positive family history of rheumatic diseases.

## Patients and Methods

### Ethics Approval

This study (AJCH-44 and AJCH-50) was approved by the Dubai HealthCare Authority Research Ethics Committee. Since this is a retrospective de-identified cohort description study, the ethics committee determined that an informed consent was not required.

### Study Design

The period of this study was from November 2016 to August 2020, and the cohort included patients with suspected or established rheumatology diseases seen as outpatient or inpatient at Al Jalila Children’s Specialty Hospital and for whom genetic testing was ordered. Prior to October 2019, most tests were referred to external CAP-accredited laboratories (n=50). Subsequently, the remaining tests (n=20) were performed in the AJCH CAP-accredited genomics laboratory which was established in 2019. In-house tests were performed according to the protocols described below.

Genetic testing included *MEFV* genotyping or sequencing (n=16), targeted NGS gene panel testing (n=33), and whole exome sequencing (WES) (n=20). Rapid trio/quad WES was performed on three cases in the Intensive Care Unit (ICU) setting. *UGT1A1* dinucleotide promoter expansion testing was performed for one patient (#33, **Table 1**), who was on methotrexate, and exhibited elevated bilirubin levels. *UGT1A1* testing had a direct impact on this patient’s management plan since it confirmed Gilbert syndrome diagnosis and rule out possible evolving hepatic toxicity due to methotrexate. A list of the NGS panels and their gene content is available in **Supplementary Methods**.

**Table 1.**
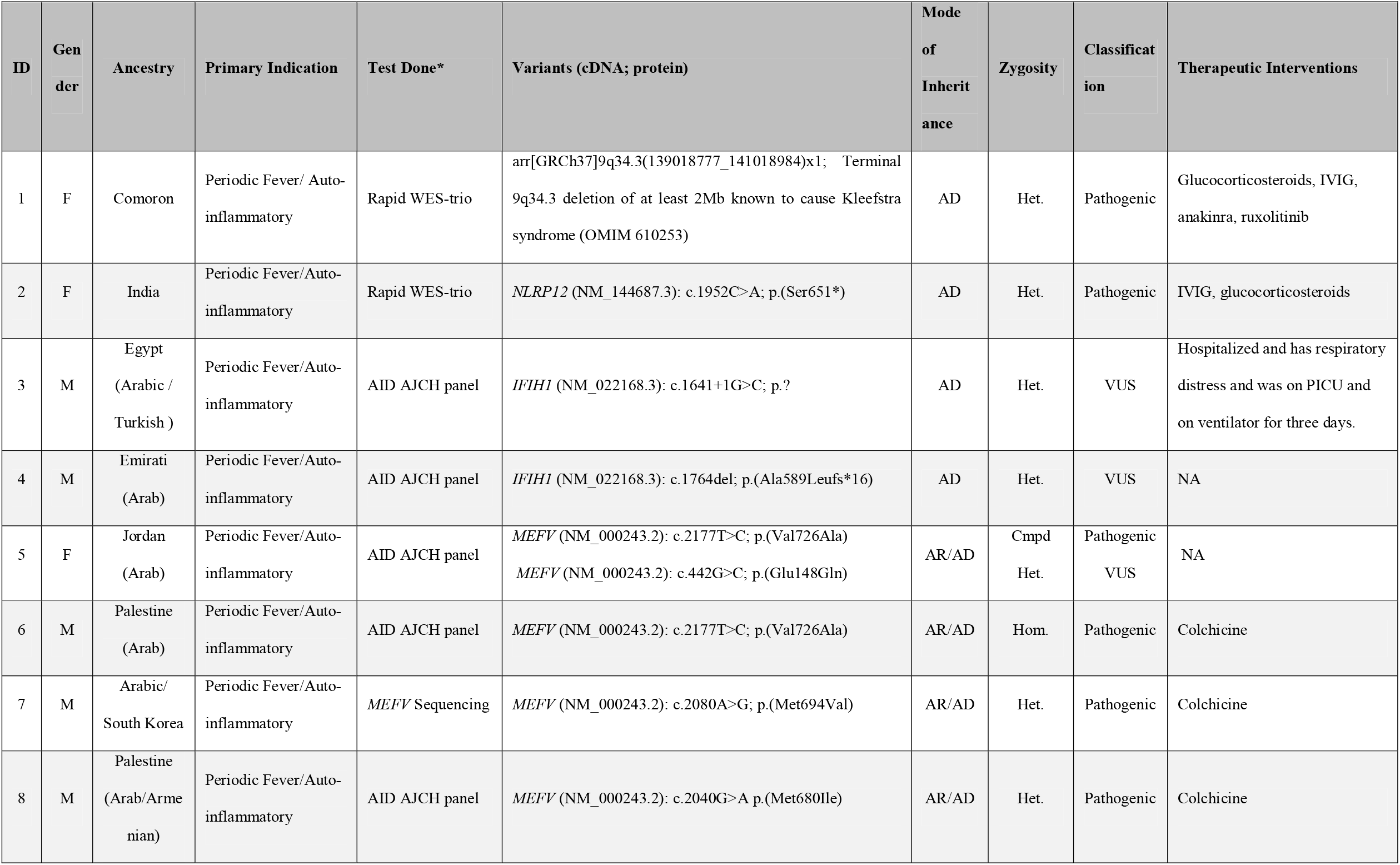
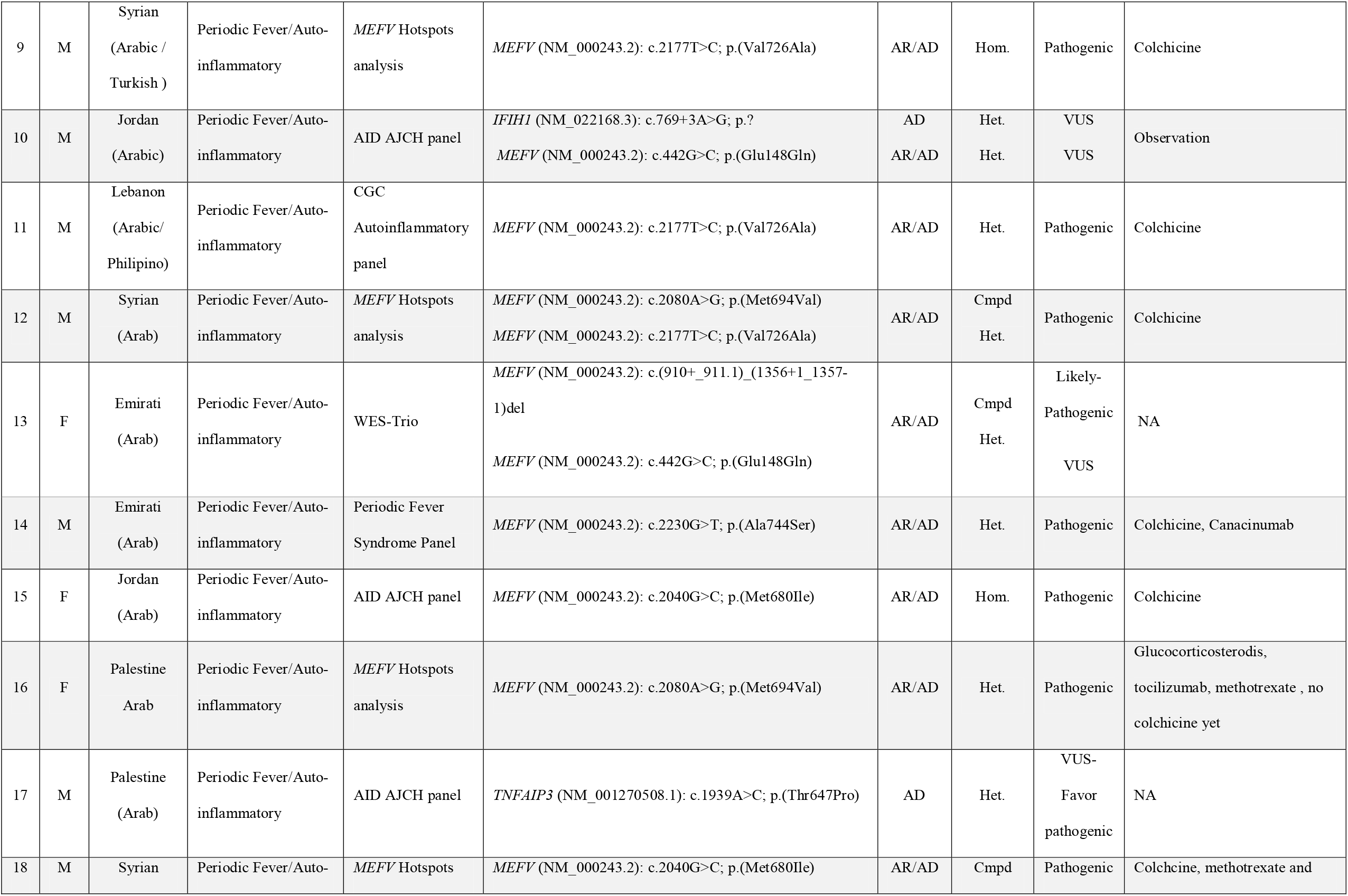
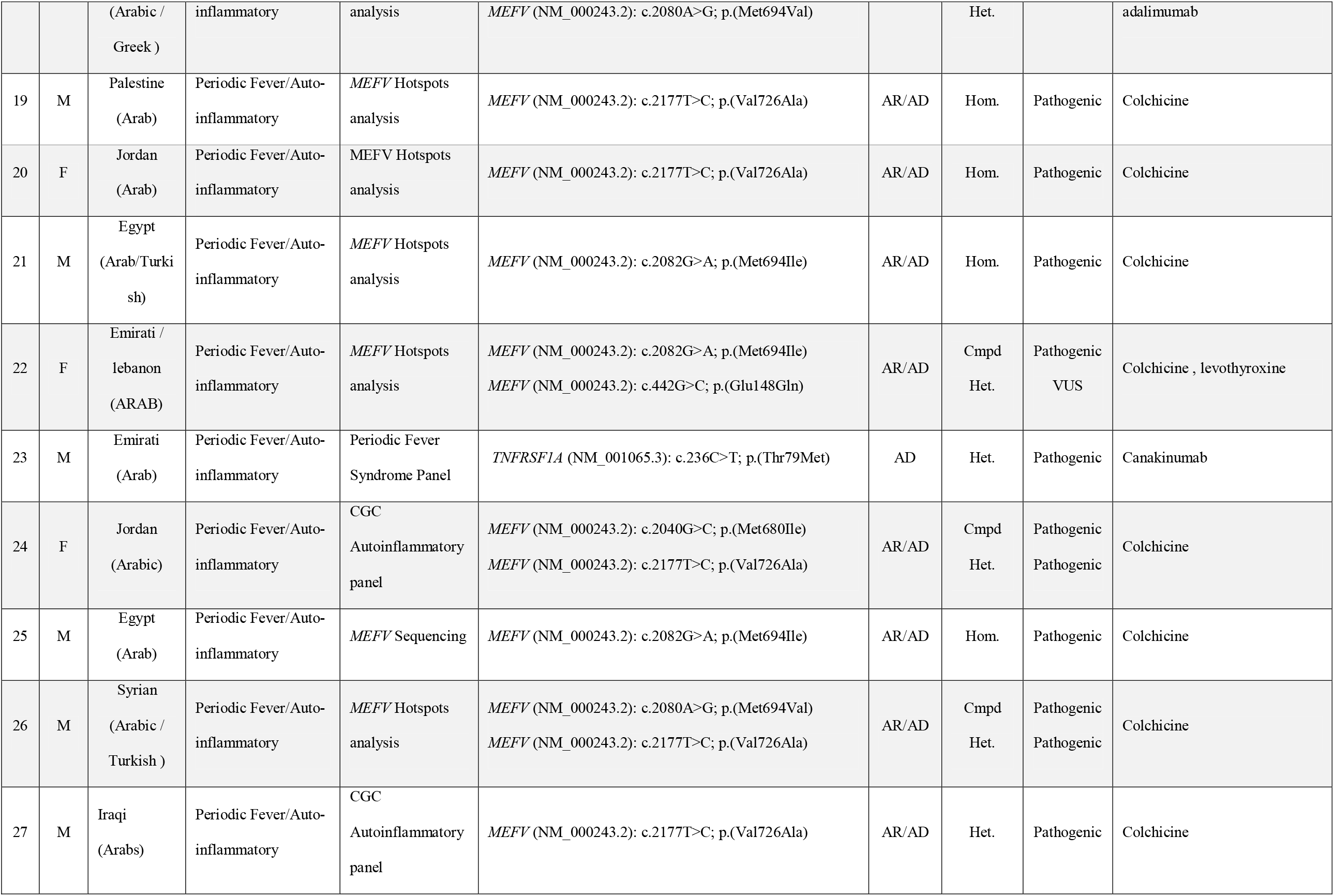
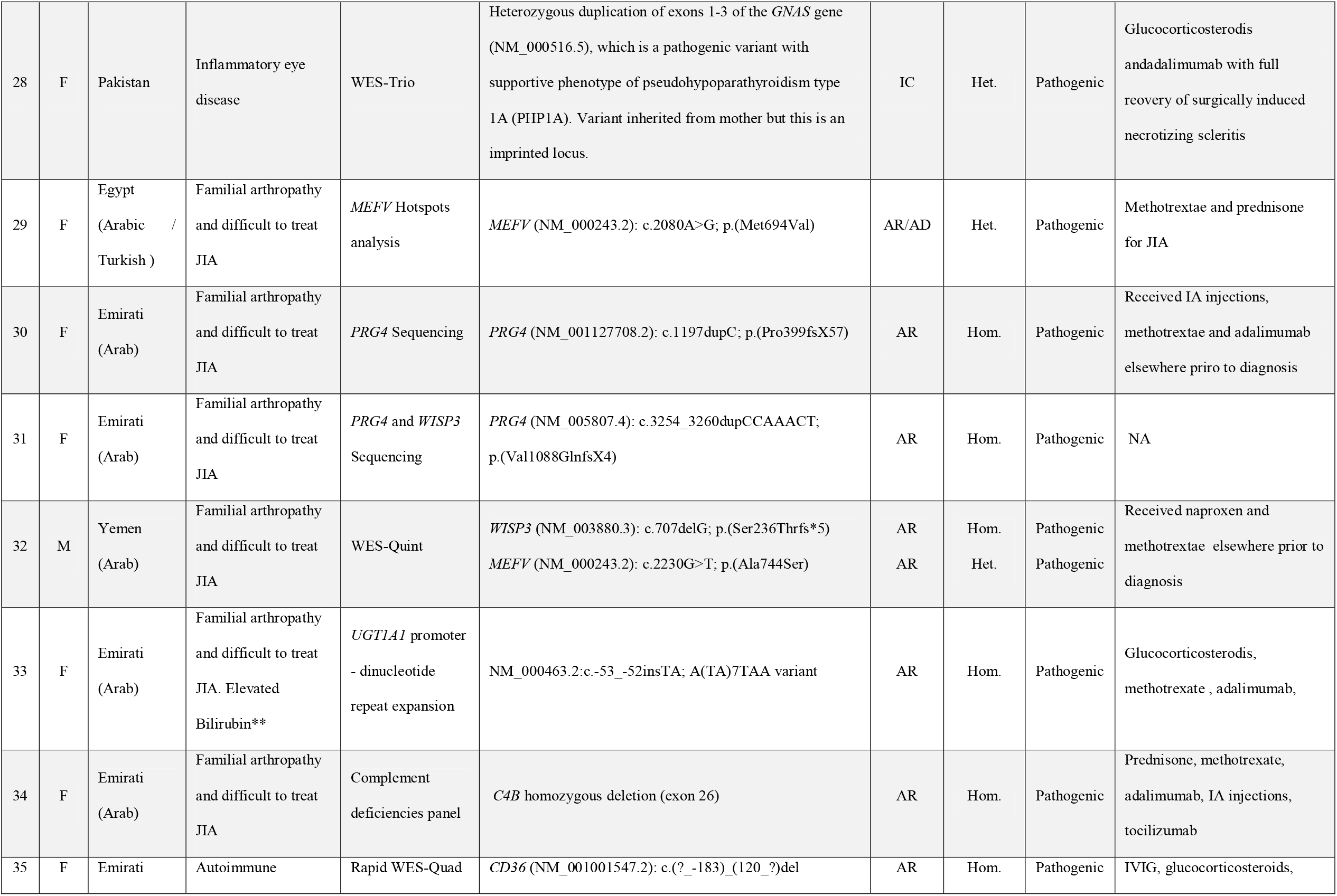
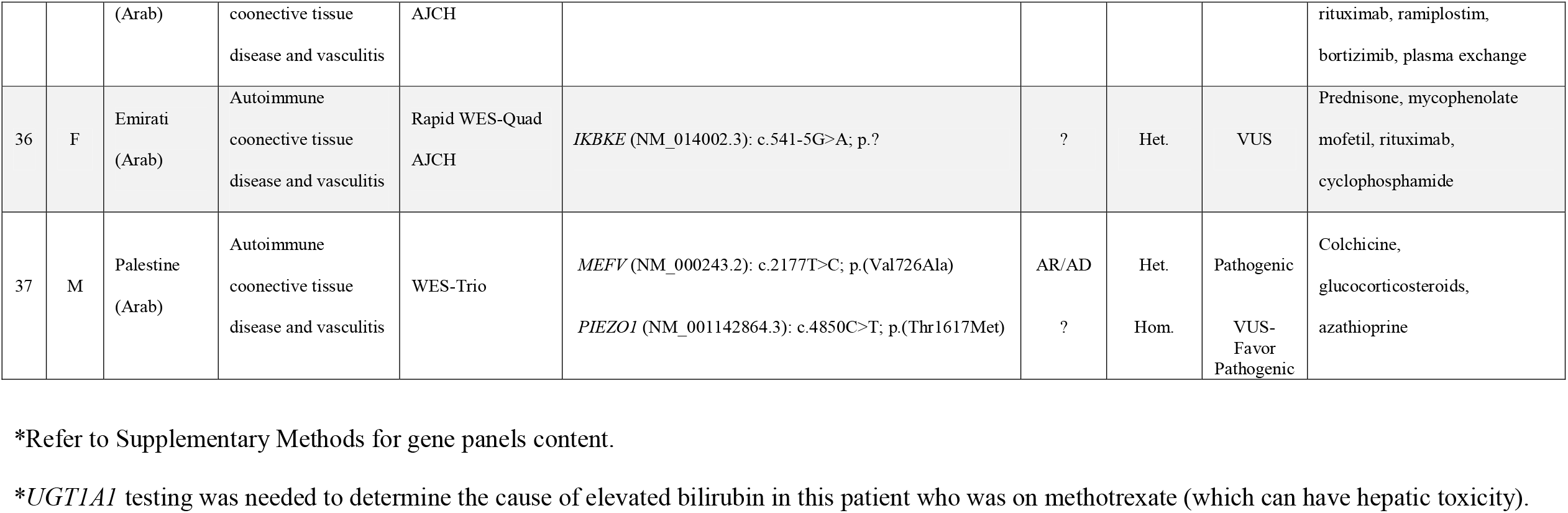
List of cases with positive or candidate findings

### Next Generation Sequencing and Whole Exome Sequencing

Genomic DNA was extracted from peripheral blood cells using standard DNA extraction protocols (Qiagen, Germany). Following fragmentation by ultra-sonication (Covaris, USA), genomic DNA was processed to generate sequencing-ready libraries of short fragments (300-400bp) using the SureSelect^XT^ kit (Agilent, USA). RNA baits targeting all coding regions were used to enrich for whole exome regions using the SureSelect Clinical Research Exome V2 kit (Agilent, USA). The enriched libraries underwent next generation sequencing (2 × 150bp) using the SP flow cell and the NovaSeq platform (Illumina, USA).

### Bioinformatics analysis and variant interpretation

Sequencing data was then processed using an in-house custom made bioinformatics pipeline to retain high quality sequencing reads with at least 100X coverage across all coding regions.

High quality variants were annotated for allele frequency, protein effects, and presence or absence in disease databases, such as ClinVar and the Human Gene Mutation Database (HGMD). Only rare variants (<0.5% if novel and <1% if present in disease databases) were retained for downstream filtration and analysis.

For targeted gene analysis, only rare, known pathogenic or novel, variants in the relevant genes associated with patient’s indication were retained for interpretation. see Supplementary Methods for a list of indication-based gene panels. For whole exome sequencing (WES), all known pathogenic variants in ClinVar/HGMD and novel loss-of-function variants in disease genes were retained. In addition, segregation analysis was performed for family-based (trio, quad, quint) WES to identify dominant, *de novo*, homozygous, and compound heterozygous variants. Finally, variants relevant to patients’ primary indications were retained for final classification and reporting. When identified, secondary findings, as recommended by the American College of Medical Genetics and Genomics (ACMGG)^8^, were returned back for patients who opted in for such findings.

All retained variants from panels and exomes were classified and reported as previously described^9,10^ using the American College of Medical Genetics and Genomics (ACMGG) sequence variant interpretation guideline^11^.

### Copy number analysis

Copy number variants (CNVs) were called using normalized NGS read depth data in the referral laboratories (patients # 1, 13, and 28) or in-house (patients # 34 and 35) using previously described algorithms^9,12,13^. Copy number changes were confirmed by microarrays (9q34.3 deletion in patient #1), specific multiplex ligation-dependent probe amplification (MLPA) (*MEFV* exons 3 – 4 deletion in patient #13 and *GNAS* exons 1 – 3 in patient #28), PCR and gel electrophoresis (*CD36* exons 2 – 3 deletion in patient #35), or a customized droplet digital PCR (ddPCR) (*C4B* exon 4 in patient #34).

### Droplet digital PCR (ddPCR) and the *C4B* assay

Droplet digital PCR (ddPCR) was performed as previously described^12,13^. Given the sequence homology in exons 26 of the *C4A* and *C4B* genes, a specific assay (C4BEx26) was designed and optimized to target unique nucleotides in the *C4B* gene. A non-specific assay (C4ABEx26) targeting both genes was also used for quality control. Target probes were FAM-labelled, while the reference *RPP30* probe was HEX-labelled. Data analysis was performed by the QuantaSoft software (Bio-Rad, USA), and the copy number state at each of the *C4A* and/or *C4B* targets was computed based on the ratio of the concentration of the target (FAM) over the reference (HEX).

The primers and probes for the specific (C4BEx26) and non-specific (C4ABEx26) assays are listed below:

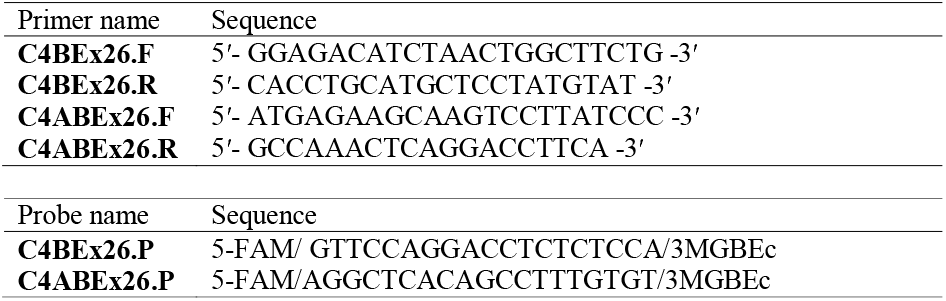

### *IFIH1* enrichment analysis

Allele counts of loss of function (LoF) variants, defined as nonsense, frameshift, or splice site ±1,2, were queried from the Genome Aggregation Database (gnomAD)^14^ or the Greater Middle East (GME)^15^ variome database. The cumulative LoF frequency in gnomAD or GME was determined as the sum of all LoF allele counts divided by the maximum total allele count in each database. This cumulative allele frequency was then compared with that in our patient cohort where the *IFIH1* gene was fully sequenced. For this analysis, the c.769+3A>G variant in patient #10 was presumed to negatively impact splicing given it affects a highly conserved nucleotide and was predicted by several *in silico* tools to alter splicing.

## Results and Discussion

### Cohort Description

Our cohort consisted of 71 pediatric (median age: 8.5 years, range: 0.88 – 18 years) patients (48% females, 52% males) who underwent genetic testing as part of work up for periodic fever/auto-inflammatory diseases (n=45), autoimmune connective tissue disorders and vasculitis (n=11), inflammatory eye diseases (n=6), and familial arthropathy or difficult to treat JIA (n=9). Most (68/71 or 96%) patients were of Arab descendant, including 28 Emiratis (39%), 13 North African (Egypt, Algeria, Morocco) Arabs (18%), 7 Syrians (10%), 6 Palestinians (8%), 6 Jordanians (8%), 2 Lebanese (3%), and 2 Yemenis (3%). Consanguinity was self-reported in 30% (21/71) of the families.

### Positive Findings

The overall diagnostic yield was 46.5% where 33 out of 71 patients had definitive pathogenic variant(s) explaining all or partial aspects of their clinical presentations, and/or guiding their medical management (**Table 1** and **Figure 1**). The positive rate was highest in patients with familial arthropathy (66.7%, n=9) and lowest in patients with inflammatory eye disease (16.7%, n=6). The diagnostic yield was 47.8% and 18.2% for periodic fever/auto-inflammatory disease (n=46) and autoimmune connective tissue disease (n=11), respectively (**Figure 1**).

**Figure 1.**
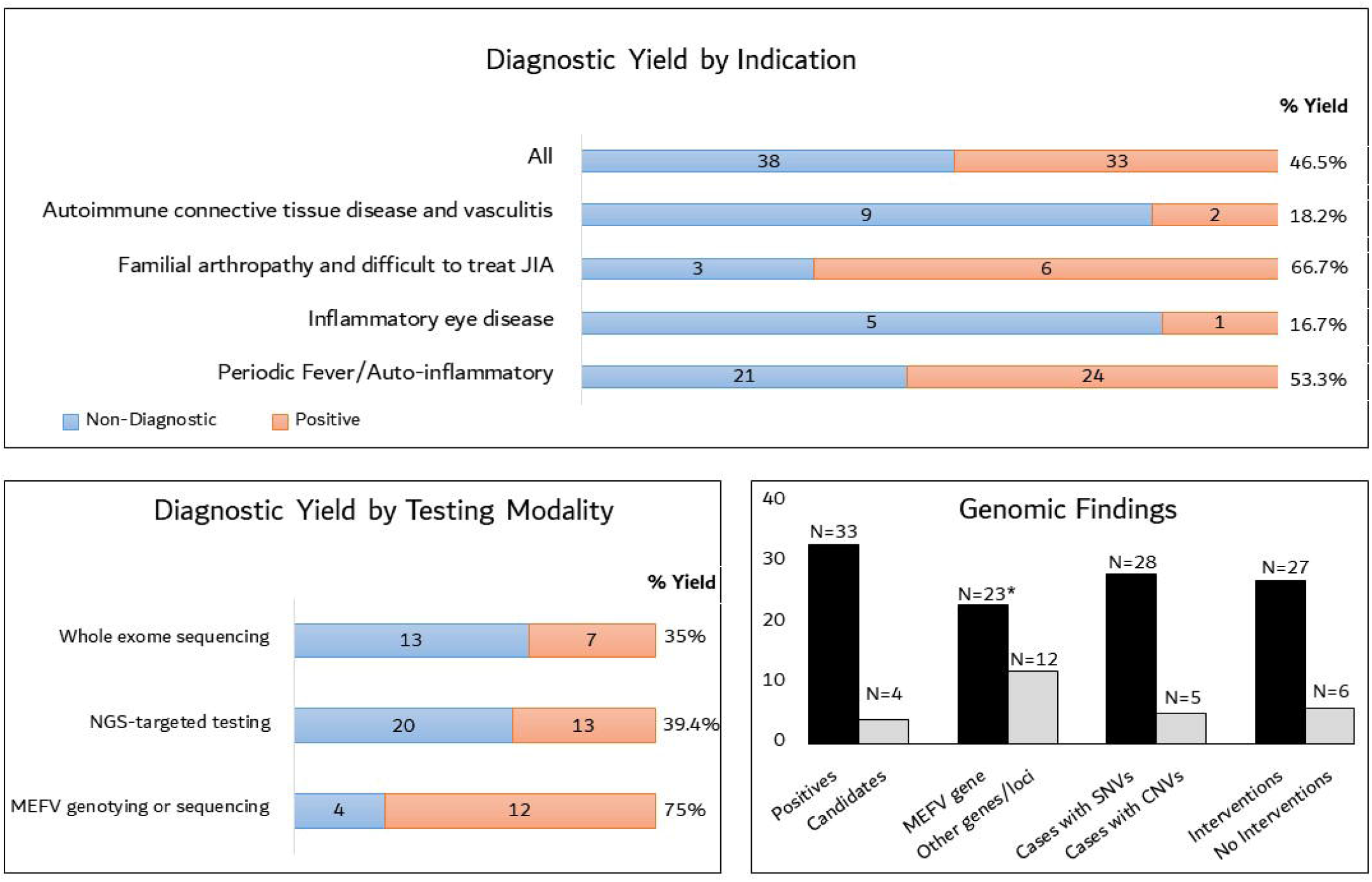
Diagnostic yield, utility and characteristic findings of genomic testing in pediatric patients with pediatric rheumatological disorders. *Top*, Diagnostic yield stratified by specific indications. *Lower left*, diagnostic yield stratified by testing modality. *Lower right*, genomic landscape of positive cases shown by numbers with pathogenic copy number variants (CNVs), single nucleotide variants (SNVs) in the *MEFV* gene or any other gene. Number of possible candidate genes/variants are shown and discussed in Results. Finally, numbers of positive cases where genetic findings patient management are shown. *Two cases with dual diagnoses in the *MEFV* gene and a second gene (*WISP3* or *PIEZO1*) are counted in each group.

Two patients (#32 and 37) had dual diagnoses where pathogenic variants in two different genes contributed to their clinical presentations and treatment plans. Patient #32 and his sibling presented with familial arthropathy and presumptive polyarticular JIA. The proband was already on naproxen and methotrexate at time of referral to our center. We suspected a form of familial noninflammatory arthropathy based on severe progressive deforming arthropathy, imaging findings including flat vertebral bodies with multiple anterior beaking, and presence of multiple affected family members. A quint exome including both patients, along with one unaffected sibling and parents identified a novel homozygous loss of function variant in *WISP3* (**Table 1**) confirming the molecular diagnosis of PPRD which not only explained the clinical phenotype but also impacted treatment decisions as PPRD has no known treatment. An unexpected p.A744S *MEFV* pathogenic variant was detected in affected members and are followed for possible evolving FMF.

Patient #37 presented with clinical and laboratory features of an autoimmune connective tissue disease suggestive of SLE including cutaneous vasculitis, arthritis, recurrent lymphedema in addition to positive serology for SLE including presence of anti-nuclear antibodies, anti-DsDNA, low C3 and C4, and presence of anti-C1q antibodies. He also had periodic episodes of fever and high inflammatory response (CRP) not typically seen in SLE. Trio WES identified a homozygous possibly pathogenic variant in *PIEZO1* explaining the recurrent lymphedema, and a heterozygous pathogenic variant in *MEFV* which explains the periodic fever and the resolution of recurrent episodes of fever when he is on colchicine.

### Mutational landscape and contribution of copy number variants

Bi-allelic or heterozygous pathogenic variants in the *MEFV* gene, known to cause autosomal dominant and recessive forms of Familial Mediterranean Fever (FMF)^16^, contributed to 23 (69%) of all positives (**Figure 1**), while the 12 positive cases (36.4%) had genetic lesions distributed between 11 genes/loci as shown in **Table 1**. Interestingly, large intragenic or multigene copy number variants (CNVs) accounted for 5 of the 33 (15.2%) positive findings. One CNV was, for the first time to our knowledge, a heterozygous two exon deletion (exons 3 and 4) in the *MEFV* gene in one patient with periodic fever (Patient #13). A homozygous deletion of exons 2 and 3 of the *CD36* gene (**Figure 2**), which can lead to an isoimmune response to CD36+ antigens via transfusion, was identified in a patient in the ICU (patient #35) with life threatening active bleeding due to chronic refractory immune thrombocytopenia explaining her refractory response to platelet transfusions. A homozygous deletion involving exon 26 of the *C4B* gene was also identified in one patient (Patient #34) with difficult to treat JIA, persistent low C4 levels, and positive family history of rheumatoid arthritis (**Table 1**). This exon has high homology to exon 26 of the *C4A* gene. To confirm our finding, we developed an allele-specific ddPCR assay to target unique nucleotides in *C4B* exon 26 (**Methods**). As shown in **Figure 3**, this homozygous deletion was specific to the *C4B* gene. Patients #1 and #28 had 9q34.3 microdeletion and a pathogenic exons 1 – 3 pathogenic duplication in *GNAS*, respectively. Both patients had complex clinical presentation with primary features consistent with auto-inflammatory disease (patient #1) and a vision threatening surgically induced inflammatory eye disease treated successfully with adalimumab (patient #28), highlighting the utility of comprehensive genomic testing in pediatric patients with complex presentations including rheumatological features.

**Figure 2.**
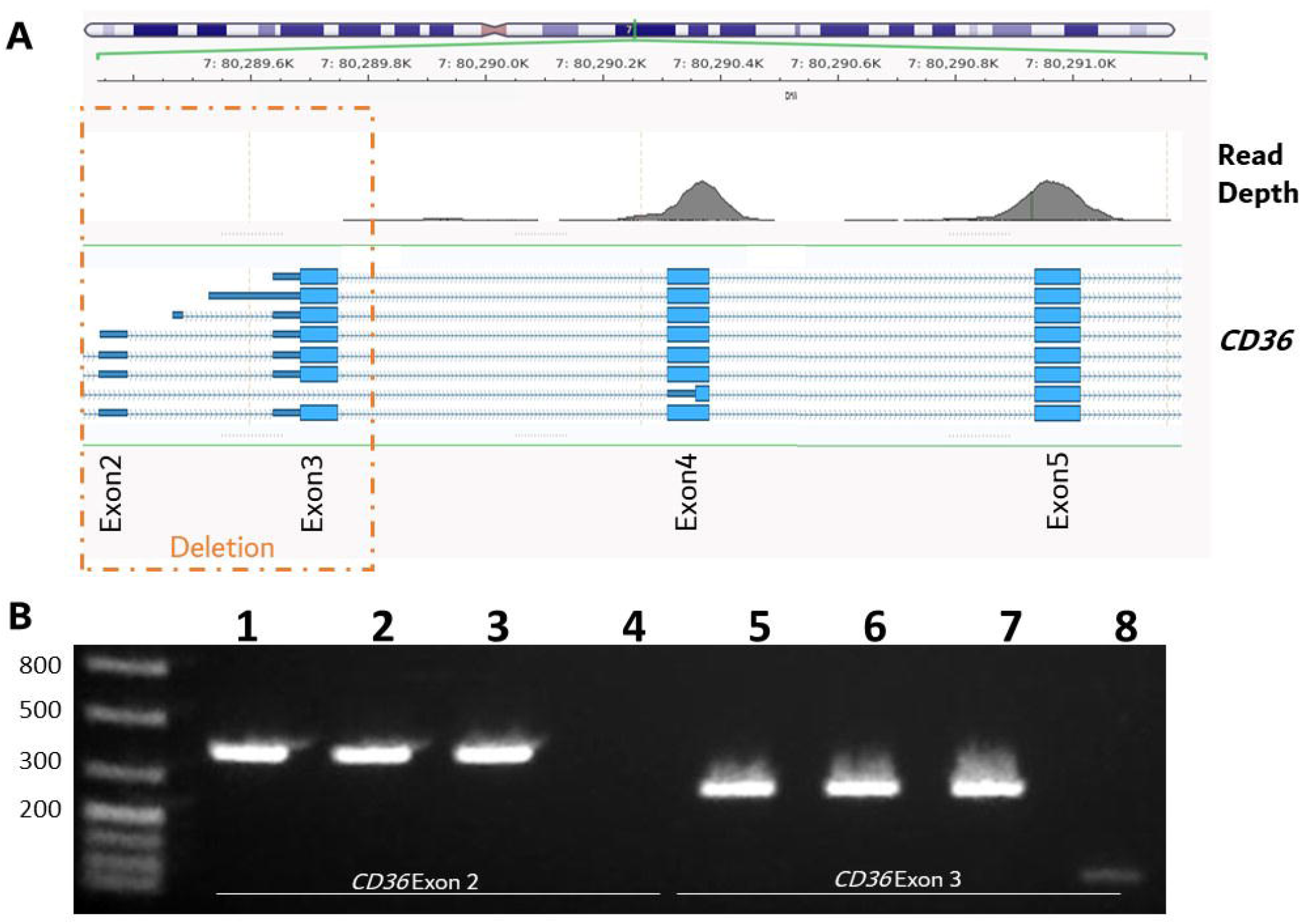
Copy number analysis of *CD36* in patient #35. **A**, NGS-based copy number analysis shows lack of sequencing reads (read depth of zero) in exons 2 and 3 of *CD36* suggesting a homozygous deletion of this region in patient #35. **B**, PCR amplification of *CD36* exons 2 (lanes 1 – 4) and 3 (lanes 5 – 8) followed by gel electrophoresis confirmed a homozygous deletion in those two exons in patient #35 (lanes 4 and 8), but not in a control sample (lanes 1 and 5), or either parent (lanes 2 and 3 for exon 2, and lanes 6 and 7 for exon 3).

**Figure 3.**
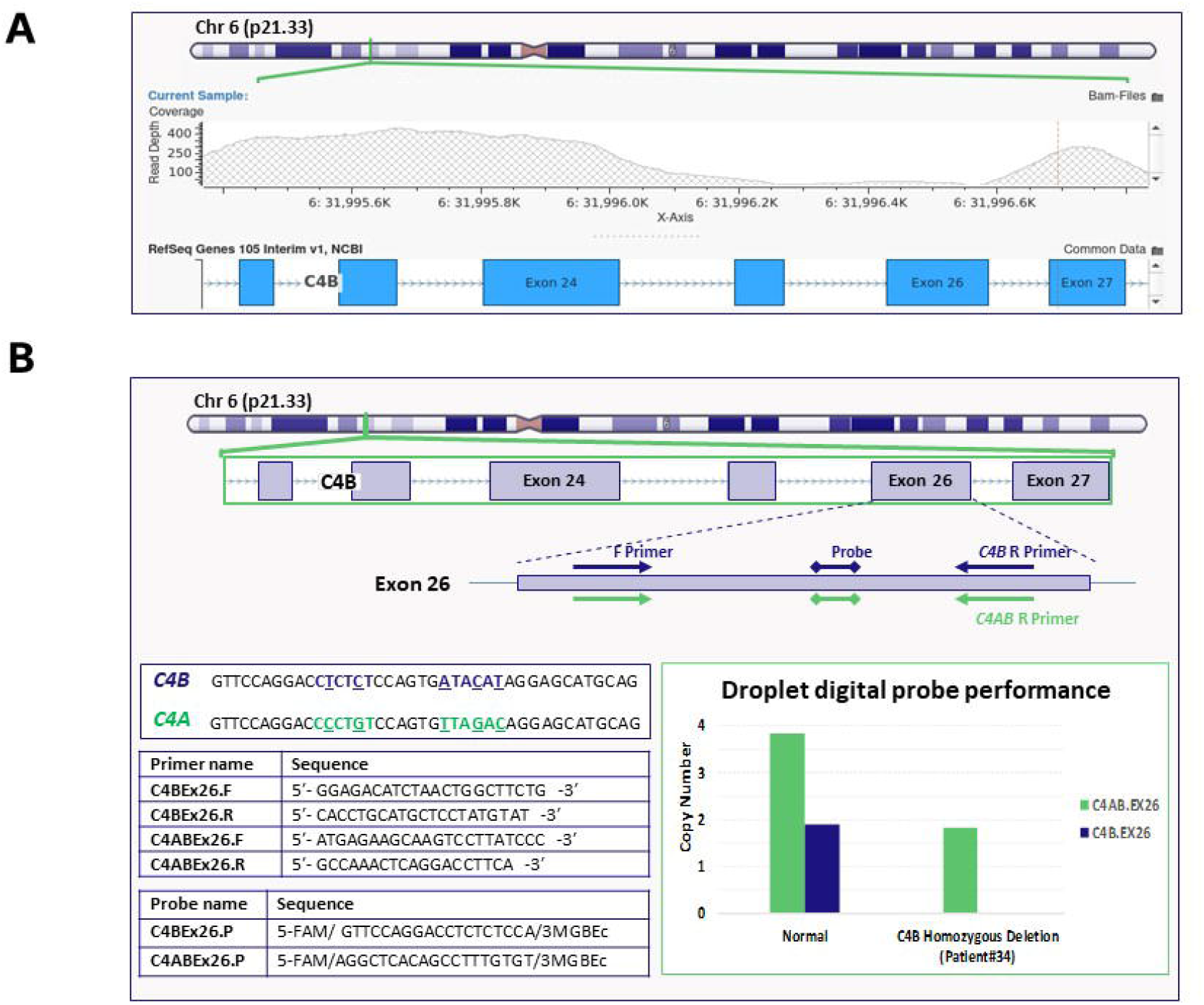
Copy number analysis of *C4B* in patient #34. **A**, NGS-based copy number analysis shows lack of sequencing reads (read depth of zero) in exons 25 and 26 of *C4B* suggesting a homozygous deletion of this region in patient #34. **B**, ddPCR confirms exon 26 homozygous deletion in patient #34. The specific assay (C4BEx26) shows zero copies of exon 26 in this patient as opposed to two copies in a control normal sample (bar graph, dark blue). On the other hand, the non-specific C4ABEx26 assay shows 4 copies (*C4A* and *C4B*) in a normal sample versus only 2 copies (*C4A* only) in patient #34 (bar graph, green). Divergent sequences in exons 26 of *C4A* (green) and *C4B* (dark blue) are highlighted. The unique sequences in *C4B* exon 26 were utilized to make specific primers/probes (listed in the tables) in this region.

### Sequencing-based diagnostics and rapid exome sequencing in urgent cases

NGS-targeted gene panels and exome sequencing had an average yield of around 37% (**Figure 1**). Interestingly, rapid exome sequencing in all 3 patients in the ICU yielded timely positive results which helped manage those patients. One patient had a terminal 9q34.3 deletion of at least 2Mb known to cause Kleefstra syndrome. Another patient has a homozygous multi-exon deletion in *CD36* gene (see above). Her sibling presenting with systemic lupus had a candidate variant in the *IKBKE* which might play a role in her clinical presentation (see below). Finally, one patient with prolonged fever, skin lesions, and a possible multisystem inflammatory syndrome had a heterozygous pathogenic loss of function variant in the *NLRP12* gene (**Table 1**).

### Candidate genes

Patients #3, 4, and #10 with auto-inflammatory disease were negative for any clinically significant variants in 36 genes, including *MEFV*, underlying the majority of causes of hereditary auto-inflammatory disease. Interestingly, the 3 patients had presumptive heterozygous LoF variants in the *IFIH1* gene, previously shown to increase susceptibility to recurrent infections/fever^17^. However, we noticed that one variant (c.1641+1G>C) was common with 1% allele frequency in the European Non Finnish population, including 6 homozygotes in gnomAD. Furthermore, the *IFIH1* gene appears to be highly tolerant to LoF variants in gnomAD. We therefore performed case enrichment analysis, and found that the cumulative frequency of LoF in our auto-inflammatory patient cohort (n=11) was 13.6% (3/22) which was significantly higher (p = 0.001; Chi-square with Yates correction) than *IFIH1* cumulative LoF frequency in *IFIH1* gene of 1.9% (1344/71996) in gnomAD. *IFIH1* cumulative LoF frequency was similarly higher (p = 0.0006; Fisher Exact Test) in our patient cohort compared to a ‘healthy’ population (n = 990) from the GME where the cumulative LoF variant frequency in *IFIH1* was 0.66% (13/1981). We therefore re-emphasize that heterozygous *IFIH1* loss of function is a common cause of recurrent infections and/or fever and should be included in genomic testing for auto-inflammatory disease. Similar to *MEFV* variants, the high allele frequency of *IFIH1* variants is not unusual for AID presentations, and diagnostic laboratories should use caution in filtering candidate variants in this gene.

Patient #36 was heterozygous for a rare splice acceptor variant in the *IKBKE* gene, previously implicated in SLE and autoimmune disease^18^. This variant is likely to contribute to this patient’s phenotype given that it is highly predicted to disrupt splicing, and that *IKBKE* is highly intolerant to loss of function variants in gnomAD. However, more functional and case level data is needed to establish a role for this gene in disease.

### Clinical Utility of Genetic Testing

Results of genetic testing had a major impact on understanding disease course and tailoring management in many patients (27 out of 33 positive cases or 82%) (**Figure 1** and **Table 1**). For example, establishing accurate diagnosis prevented use of unnecessary immune suppressive medications in patients with primary non-inflammatory arthropathy. In patient #32 and his sibling, a novel *WISP3* variant supported the diagnosis of PPRD in the proband and his sibling. Establishing accurate diagnosis was essential to understand lack of response to immune suppressive medications, for withholding methotrexate and any other immune suppressive medications, and for family counselling. This concept is also true for patient #30 and her sibling with camptodactyly-arthropathy-coxa vara-pericarditis syndrome.

Another direct impact of genetic testing is proper selection of therapeutic intervention in patients with periodic fever syndromes. This testing was essential in diagnosis of Tumor Necrosis Factor Receptor Associated Periodic Syndrome (TRAPS) in patient #23 with periodic fever. This result supported the selection of canakinumab as drug of choice whereas colchicine is the first drug of choice in patients with FMF. For some cases, there was no direct impact of test results on management, though provided relevant information as to why the disease course was inconsistent with expected natural history. For example, patient #37 with features of SLE overlapping with a periodic fever syndrome and elevated CRP typically not seen in the context of SLE. This overlap of autoimmune and autoinflammatory phenotypes can be explained in some entities such as hypocomplementemic urticarial vasculitis and, although WES was negative for this entity, he did have a heterozygous disease-causing variant in *MEFV* that could explain his recurrent fever and the responsiveness of these episodes to colchicine upon medication.

Lack of complete clinical response to conventional treatment can be explained in patient #34 with partially controlled JIA due to presence of homozygous *C4B* deletion. Furthermore, refractory chronic immune thrombocytopenia purpura (ITP) in patient #35 with no response to multiple medications was noted to have no measurable or sustained response to platelet transfusions in the ICU setting with life threatening perfuse bleeding and was found to have a homozygous *CD36* deletion. In this case, the child, who is CD36 negative, had transfusions with CD36 positive platelets theoretically leading to rapid destruction of transfused platelets.

In summary, this work highlights the utility of comprehensive genomic testing in diagnosing and managing pediatric patients with a spectrum of rheumatological disorders. It also characterizes the genomic landscape of those disorders where copy number variants, high allele frequency variants, and candidate genes, such as *IFIH1*, can be common causes of the disease.

## Data Availability

All data is available upon request

## Acknowledgement

We thank all members of the genomics Center and Al Jalila Children’s specialty hospital for contributing to the workup of patients in our study cohort.

