## Supplementary Methods for "The Genomic Landscape of Pediatric Rheumatology Disorders in the Middle East"

Fathalla, et al.

**Supplementary Methods**

**Gene content of the NGS targeted analysis in this study:**

**AJCH Auto inflammatory (AID) 37 gene panel (N = 11 patients)**

*ACP5, ADA2, ADAM17, ADAR, AP1S3, CARD14, COPA, ELANE, IFIH1, IL1RN, IL36RN, LPIN2, MEFV, MVK, NLRC4, NLRP1, NLRP12, NLRP3, NOD2, OTULIN, PLCG2, POLA1, PSMB8, PSTPIP1, RNASEH2A, RNASEH2B, RNASEH2C, SAMHD1, SH3BP2, SLC29A3, STING1, TADA2A, TNFAIP3, TNFRSF1A, TREX1, TRNT1, USP18*

**CGC Auto inflammatory panel 34 gene panel (N = 4 patients)**

*ACP5, ADAM17, ADAR, AP1S3, CARD14, CECR1, COPA, IFIH1, IL1RN, IL36RN, LPIN2, MEFV, MVK, NLRC4, NLRP1, NLRP12, NLRP3, NOD2, OTULIN, PLCG2, POLA1, PSMB8, PSTPIP1, RNASEH2A, RNASEH2B, RNASEH2C, SAMHD1, SH3BP2, SLC29A3, TMEM173, TNFAIP3, TNFRSF1A, TREX1, USP18*

**Moyamoya disease (N= 1 patient)**

*RNF213*

**Periodic Fever Syndrome 31 gene panel (N = 5 patients)**

*AP1S3, CARD14, CECR1, ELANE, FOXD3, HAX1, IL10, IL10RA, IL10RB, IL1RN, IL36RN, LPIN2, MEFV, MVK, NLRC4, NLRP1, NLRP12, NLRP3, NLRP7, NOD2, PLCG2, PSMB8, PSTPIP1, RAB27A, RBCK1, RNF31, SH3BP2, SLC29A3, TMEM173, TNFRSF11A, TNFRSF1A*

**Complement deficiencies 33 gene panel (N = 1 patient)**

*C1QA, C1QB, C1QC, C1R, C1S, C2, C3, C4A, C4B, C5, C6, C7, C8A, C8B, C8G, C9, CD46, CD59, CFB, CFD, CFH, CFHR1, CFHR2, CFHR3, CFHR4, CFHR5, CFI, CFP, FCN3, ITGAM, MASP2, SERPING1, THBD*

**Hereditary hemophagocytic lymphohistiocytosis** **(HLH) 10 gene Panel (N = 2 patients)**

*AP3B2, BLOC1S6, PRF1, UNC13D, STX11, STXBP2, RAB27A, LYST, SH2D1A, XIAP*

**Rhabdomyolsis 91 gene panel (N = 1 patient)**

*ACAD9, ACADM, ACADVL, ACTA1, AGL, ALDOA, ANO5, B3GALNT2, B4GAT1, BAG3, CAPN3, CASQ1, CPT1A, CPT2, CRYAB, CTDP1, DAG1, DGUOK, DMD, DNA2, DNAJB6, DPM1, DPM2, DYSF, EMD, ENO3, ETFA, ETFB, ETFDH, FHL1, FKRP, FLAD1, FLNC, G6PD, GAA, GBE1, GMPPB, GNE, GYG1, GYS1, HADHA, HADHB, HMBS, ISCU, ISPD, ITGA7, KBTBD13, KLHL40, LAMA2, LAMP2, LDB3, LDHA, LMNA, LPIN1, MEGF10, MICU1, MTM1, MYOT, NEB, PFKM, PGAM2, PGK1, PGM1, PHKA1, PHKB, PNPLA2, POMGNT1, POMT1, POMT2, PYGM, RBCK1, RRM2B, RYR1, SGCA, SGCB, SGCD, SGCG, SIL1, SLC22A5, SLC25A20, SUCLA2, SUCLG1, TANGO2, TCAP, TK2, TNNT1, TNPO3, TPM2, TPM3, TRIM32, FDX2.*

**Blau syndrome (N= 4 patients)**

*NOD2*

**Osteogenesis imprefecta AJCH 48 gene panel (N= 1 patient)**

*COL1A1, COL1A2, BMP1, CRTAP, FKBP10, P3H1, PLOD2, PPIB, SEC24D, SERPINH1, SPARC, TMEM38B, CREB3L1, IFITM5, MBTPS2, MESD3, SERPINF1, SP7, TENT5A, WNT1, LEPRE1, FAM64A, PLS3, ALPL, ANO5, B3GAT3, B4GALT7, CLCN5, COL1A1, DMP1, FGF23, ENPP1, GNAS, GORAB, LMNA, LRP5, MAFB, MMP2, NBAS, NOTCH2, P4HB, PHEX, PLOD3, SLC34A3, TAPT1, TNFRSF11A, TNFRSF11B, XYLT2.*

**Arthropathy panel (N= 2 patients;** *one patient had *PRG4* sequencing only)

*PRG4, WISP3*

**ADA2-related disease (N= 1 patient)**

*ADA2*
